# Neutralizing antibody response in non-hospitalized SARS-CoV-2 patients

**DOI:** 10.1101/2020.08.07.20169961

**Authors:** Natalia Ruetalo, Ramona Businger, Karina Althaus, Simon Fink, Felix Ruoff, Klaus Hamprecht, Bertram Flehmig, Tamam Bakchoul, Markus F. Templin, Michael Schindler

## Abstract

The majority of infections with SARS-CoV-2 are asymptomatic or mild without the necessity of hospitalization. It is of importance to reveal if these patients develop an antibody response against SARS-CoV-2 and to define which antibodies confer virus neutralization. We conducted a comprehensive serological survey of 49 patients with a mild course of disease and quantified neutralizing antibody responses against a clinical SARS-CoV-2 isolate employing human cells as targets.

Four patients (8%), even though symptomatic, did not develop antibodies against SARS-CoV-2 and two other patients (4%) were only positive in one of the six serological assays employed. For the remainder, antibody response against the S-protein correlated with serum neutralization whereas antibodies against the nucleocapsid were poor predictors of virus neutralization. Regarding neutralization, only six patients (12%) could be classified as highly neutralizers. Furthermore, sera from several individuals with fairly high antibody levels had only poor neutralizing activity. In addition, employing a novel serological Western blot system to characterize antibody responses against seasonal coronaviruses, we found that antibodies against the seasonal coronavirus 229E might contribute to SARS-CoV-2 neutralization.

Altogether, we show that there is a wide breadth of antibody responses against SARS-CoV-2 in patients that differentially correlate with virus neutralization. This highlights the difficulty to define reliable surrogate markers for immunity against SARS-CoV-2.

## Introduction

The most recent emerging virus outbreak happened in China in December 2019 caused by SARS (severe acute respiratory syndrome) coronavirus-2 (SARS-CoV-2) (1), leading to a pandemic, as defined by the WHO in March 2020 (2, 3). Infections with SARS-CoV-2 can cause the so-called disease COVID-19 (Coronavirus disease 2019). 50% of all COVID-19 cases range from asymptomatic to mild. 30% show moderate to pronounced symptoms. 5-20% of patients are hospitalized due to critical course of infection with severe lung complications and on average ~5% die, even though there is high variation dependent on the country (4). Recent data from a multicentric cohort of 10021 hospitalized COVID-19 patients, showed an in-hospital mortality of 73% in mechanically ventilated patients requiring dialysis, and of 53% of invasively ventilated patients (5). One critical determinant of illness is age, as mortality is highest in the elderly population (1, 2, 4, 5). SARS-CoV-2 is currently spreading in an immune naive population and a vaccine is not yet available, even though there are numerous candidates in the advanced development pipeline (6). For an updated online resource refer to https://biorender.com/covid-vaccine-tracker.

The pandemic is not only devastating in terms of the direct harm to human health inflicted by the virus infection, but the continuous quarantine and lock-down measurements have enormous negative impact on the socio-economical life of billions of individuals (7). With the potential of achieving herd immunity as the virus spreads within the population, numerous studies analyzing the prevalence of SARS-CoV-2-specific antibodies in the population have been initiated (8-11).

Regardless of the prevalence of antibodies against SARS-CoV-2, one still poorly defined determinant is what type of antibodies neutralize SARS-CoV-2 and hence potentially confer protective immunity against the infection, even though very recent data using Vero cells and pseudovirus systems suggest IgG against the receptor binding domain play a role (12, 13). Besides, antibodies that bind to SARS-CoV-2 but do not result in neutralization might enhance infection, a phenomenon called antibody-dependent enhancement (ADE) (14, 15), which has not been investigated either. Finally, the role of cross-protecting antibodies from seasonal coronaviruses is also discussed, but not yet experimentally assessed (14).

To shed further light on the determinants of human serum to neutralize SARS-CoV-2 we performed a comprehensive serological analysis of 49 individuals that were non-hospitalized and range from an asymptomatic to a mild course of disease. We employed several assays measuring SARS-CoV-2-specific IgGs against the S-protein, the S-protein RBD, and the nucleocapsid. Furthermore, we assessed S-RBD specific IgM and IgA and used a novel innovative throughput Western-blot system to detect IgGs against SARS-CoV-2 and seasonal coronaviruses. Finally, all serological parameters were associated with the ability of the 49 sera to neutralize the infection on human cells with a clinical SARS-CoV-2 isolate.

## Results

### The majority of patients develop SARS-CoV-2-specific antibodies

For our serological survey, we recruited individuals coming to the Department of Transfusion Medicine to donate blood for plasma therapy. All 49 patients included in this study were non-hospitalized with asymptomatic to mild courses of disease, including cough (69%), fever (59%), limb pain and headache (35%), diarrhea (10%), and loss of taste (10%) (Supplementary Table 1). The age ranged from 19-66 years (median 40 years), gender was balanced (24 male, 25 female). The time from positive SARS-CoV-2 test to blood sampling was 14-64 days (median 45 days).

We employed several serological assays to detect antibodies against SARS-CoV-2 (Table 1 and Supplementary Table 1). IgG ELISAs against the S-protein (Euroimmune) and S-protein RBD (Mediagnost), IgA and IgM against S-RBD (Mediagnost) as well as an ECLIA detecting IgG against the viral nucleocapsid (NC, Roche). Furthermore, we applied a throughput Western blot system (DigiWest) allowing detection of SARS-CoV-2 and seasonal coronavirus antibodies (16).

**Table 1.** Serological parameters of sera and VNT_50_ vlaues.

| Patient ID | IgG S | IgG S-RBD | IgG NC | SCoV-2 IgG | VNT50 |
| --- | --- | --- | --- | --- | --- |
| S11 | - | - | - | - | <20 |
| S9 | - | - | - | - | <20 |
| S48 | - | - | - | - | <20 |
| S2 | - | - | - | + | <20 |
| S25 | + | + | + | + | 20 |
| S47 | + |  | + | + | 42,9 |
| S10 | + | + | - | + | 107,1 |
| S41 | + | + | + | + | 139,6 |
| S12 | + |  | - | + | 143,7 |
| S46 | + |  | - | - | 148,5 |
| S7 |  | - | + | + | 148,9 |
| S35 | + | + | + | + | 153,7 |
| S37 | + | + | + | + | 156,5 |
| S8 | + | + | + | + | 160,4 |
| S49 | + | + | + | + | 162,1 |
| S40 | + |  | - | + | 164,4 |
| S22 | + | + | + | + | 208,6 |
| S34 | + | + | + | + | 218,7 |
| S6 | - | - | - | - | 219,4 |
| S44 | + |  | + | + | 225,2 |
| S1 | + | + | - | + | 231,8 |
| S26 | + | + | + | + | 238,2 |
| S21 | + | + | + | + | 246,9 |
| S32 | + | + | + | + | 251,2 |
| S29 | + | + | + | + | 265,8 |
| S45 | + | + | + | + | 278,1 |
| S24 | + | + | + | + | 315,5 |
| S30 | + | + | + | + | 345,3 |
| S38 | + |  | + | + | 360,9 |
| S39 | + | + | + | + | 361,5 |
| S3 | + | + | + | + | 466 |
| S33 | + | + | + | + | 489,7 |
| S20 | + | + | + | + | 490,9 |
| S43 | + | + | + | + | 521,6 |
| S14 | + | + | + | + | 542,9 |
| S28 | + | + | + | + | 579 |
| S42 | + | + | + | + | 597,7 |
| S27 | + | + | + | + | 605,7 |
| S17 | + | + | + | + | 612,7 |
| S36 | + | + | + | + | 616,1 |
| S18 | + | + | + | + | 663,6 |
| S5 | + | + | + | + | 663,6 |
| S4 | + | + | + | + | 846,7 |
| S31 | + | + | + | + | 1025 |
| S15 | + | + | + | + | 1157,1 |
| S19 | + | + | + | + | 1373,4 |
| S23 | + | + | + | + | 1447,8 |
| S13 | + | + | + | + | 2119,5 |
| S16 | + | + | + | + | 2560 |
| HP_T1 | n.a. | + | + | + | 2560 |
| HP_T2 | n.a. | + | + | + | 1454,8 |
| HD1 | n.a. | - | - | - | <20 |
| HD2 | n.a. | - | - | - | <20 |
| HD3 | n.a. | - | - | - | <20 |
| HD4 | n.a. | - | - | - | <20 |

4/49 (8%) sera were negative in all serological assays employed to detect SARS-CoV-2-specific antibodies, even though they were symptomatic showing two or more symptoms. In addition, two more sera were only positive in one of the four assay systems to detect IgGs against S or NC, which is the reason why we consider these sera also negative. Apart from the robust IgG-response both against the S-protein (90%) and NC (80%), development of S-specific IgM and IgA was less prominent, with 35 and 30% of positive sera respectively (Supplementary Table 1).

In sum, even though 12% (6/49) of patients did not develop antibodies against SARS-CoV-2, the vast majority (88%, 43/49) of individuals mount a robust SARS-CoV-2-specific antibody response.

### Few patients develop high virus neutralizing titers (VNTs) after SARS-CoV-2 infection

The majority of SARS-CoV-2 infected individuals seroconvert within 14 days (Guo et al., 2020); however, it is less clear how potently sera from these patients neutralizes SARS-CoV-2 (17). To test for virus neutralization we established two procedures using human Caco-2 cells as targets (Fig. 1a). First, we infected cells with a SARS-CoV-2 strain isolated from a throat swab of a patient showing a high viral load as determined by qRT-PCR. This strain was designated SARS-CoV-2 200325_Tü1. Cells were co-incubated with patient sera and virus in serial two-fold dilutions from 1:20 up to 1:2560. 48 hpi cells were fixed with 80% acetone and immunofluorescence stained against SARS-CoV-2 antigens with a highly potent patient serum we retrieved from a hospitalized convalescent donor. Cells were counter-stained with DAPI and infection rates quantified via automated fluorescence-microscopy. For the second approach, we employed the mNeonGreen expressing infectious SARS-CoV-2 clone [12]. Cells were treated and infected exactly as explained for the SARS-CoV-2 Tü1 strain, but using slightly adjusted dilutions of sera; 1:40 to 1:5120. 48 hpi cells were fixed with 2% PFA containing Hoechst33342 as nuclear stain (Fig. 1b, representative sera examples of both procedures). Infection rates of the corresponding serum dilutions were used to plot sigmoidal inhibition curves and calculate the virus-neutralizing titer 50 (VNT_50_) which is the serum dilution inhibiting the half-maximal infection (Fig. 1c). The VNT_50_ values of the sera obtained with the primary patient isolate correlated highly significantly with the titers calculated when using the mNeonGreen-expressing infectious clone (r=0.7349; Fig. 1d). We only obtained slight discrepancies for highly potent sera that seemed to neutralize SARS-CoV-2-mNG more efficiently than SARS-CoV-2-Tü1 at high dilutions. In both assays, sera from four healthy donors and two consecutive sera from a hospitalized convalescent patient were either completely negative for VNT as well as all serological assays, or showed robust neutralization and SARS-CoV-2-specific antibodies (Table 1).

**Figure 1.**
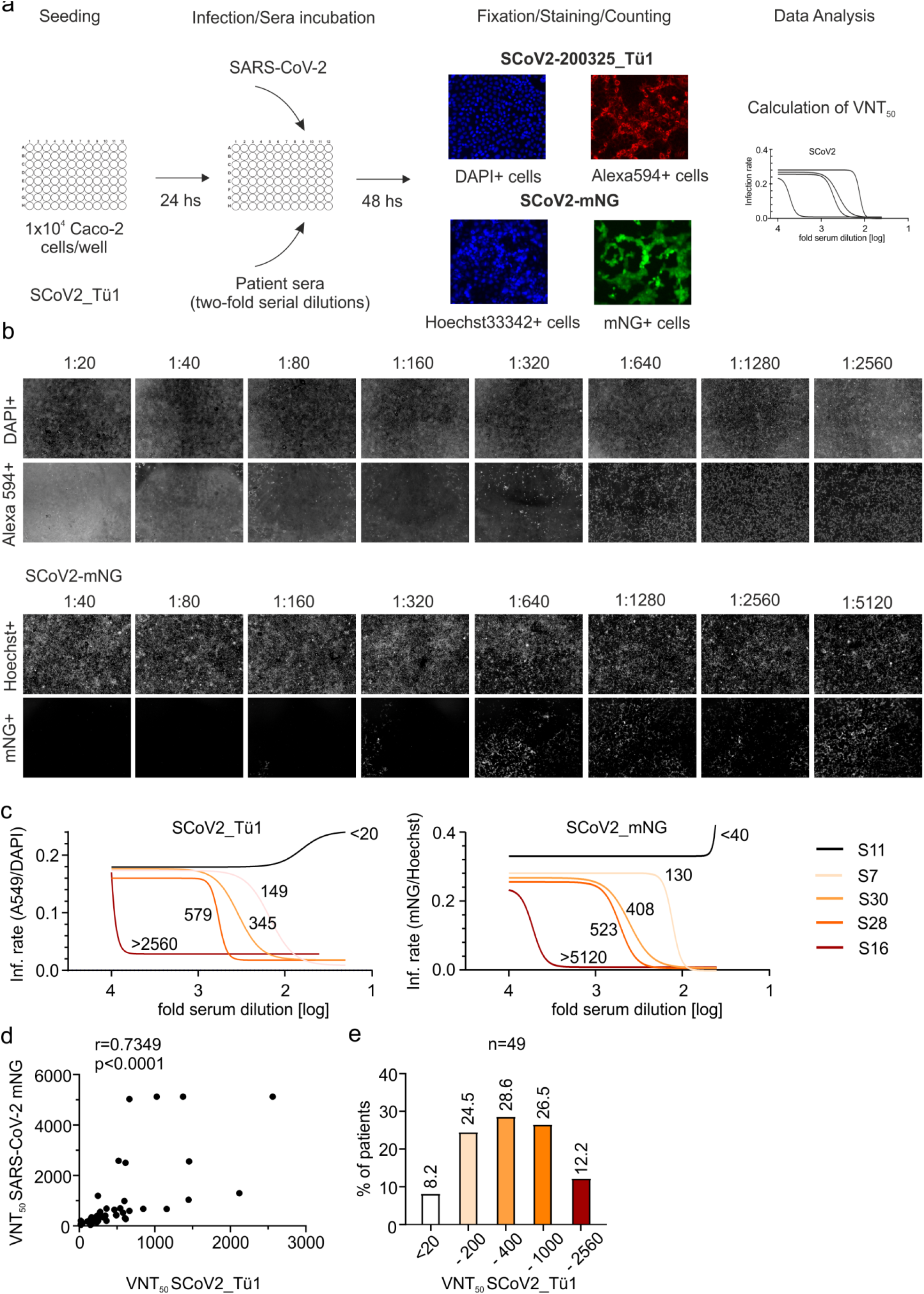
Neutralization of SARS-CoV-2 by sera of COVID-19 convalescent patients. (a) Experimental layout of the two neutralization assays employed using the clinical isolate (SARS-CoV-2_Tü1) and the green-fluorescent virus (SARS-CoV-2_mNG). (b) Primary data showing results of both neutralization assays using one patient serum as an example (S28). In the upper row, the total amount of cells for each well of the two-fold serial dilution of sera is shown, as DAPI+/Hoechst+ respectively. In the lower, infected cells are visualized, indicated as Alexa594+/mNG+ cells, respectively. (c) Neutralization curves of five representative sera measured by both assays. The graphs show the nonlinear regression fitting calculated for five patients who displayed different neutralization capacity: no, poor, low, medium, and high neutralization. The VNT_50_ for each patient are shown next to each curve. (d) Correlation analysis of VNT_50_ measured by both assays (n=49). (d) Percentage of patients classified according to the VNT_50_ using SARS-CoV-2_Tü1. The titers used to classify the sera are shown below the columns: <20, 20-200, 201-400, 401-1000 and 1000-2560. Above the columns is shown the percentage of sera that correspond to each category.

Patient sera were classified according to their neutralizing capacity (Fig. 1e) revealing that only 12% (6/49) were highly potent neutralizers. 8% (4/49) of sera did not neutralize

SARS-CoV-2 at all *in vitro* and 24% (12/49) were poor neutralizers even though showing robust signals in the different serological assays employed. Of note, using human Caco-2 cells that express human Fc-receptors (18), we should be able to observe ADE. However, none of the sera enhanced infection of SARS-CoV-2 at any dilution, arguing against ADE, at least in our system. Hence, there is a large diversity in the ability of patient sera to neutralize SARS-CoV-2 which is not always associated with the amount of SARS-CoV-2-specific antibodies.

### VNT_50_ is not associated with specific patient characteristics, apart from gender

We next assessed potential associations of patient characteristics with serum neutralization. Neither the sampling date 14-64 days (Fig. 2a) nor patient age (Fig. 2b) correlated with serum neutralization. This indicates that seroconversion, as reported, is achieved within 14 days in all patients and VNTs might not drop up to 64 days. On average, titers were higher in males as compared to females (Fig. 2c). In detail, 3/4 individuals that did not neutralize at all were female but 5/6 highly potent neutralizers were male (Supplementary Table 1). Furthermore, the average VNT_50_ of men is double as compared to women (613 vs 322; compare Fig. 2c). Of note, there was no association of VNT_50_ with the severity of disease (Fig. 2d), respectively amount of symptoms.

**Figure 2.**
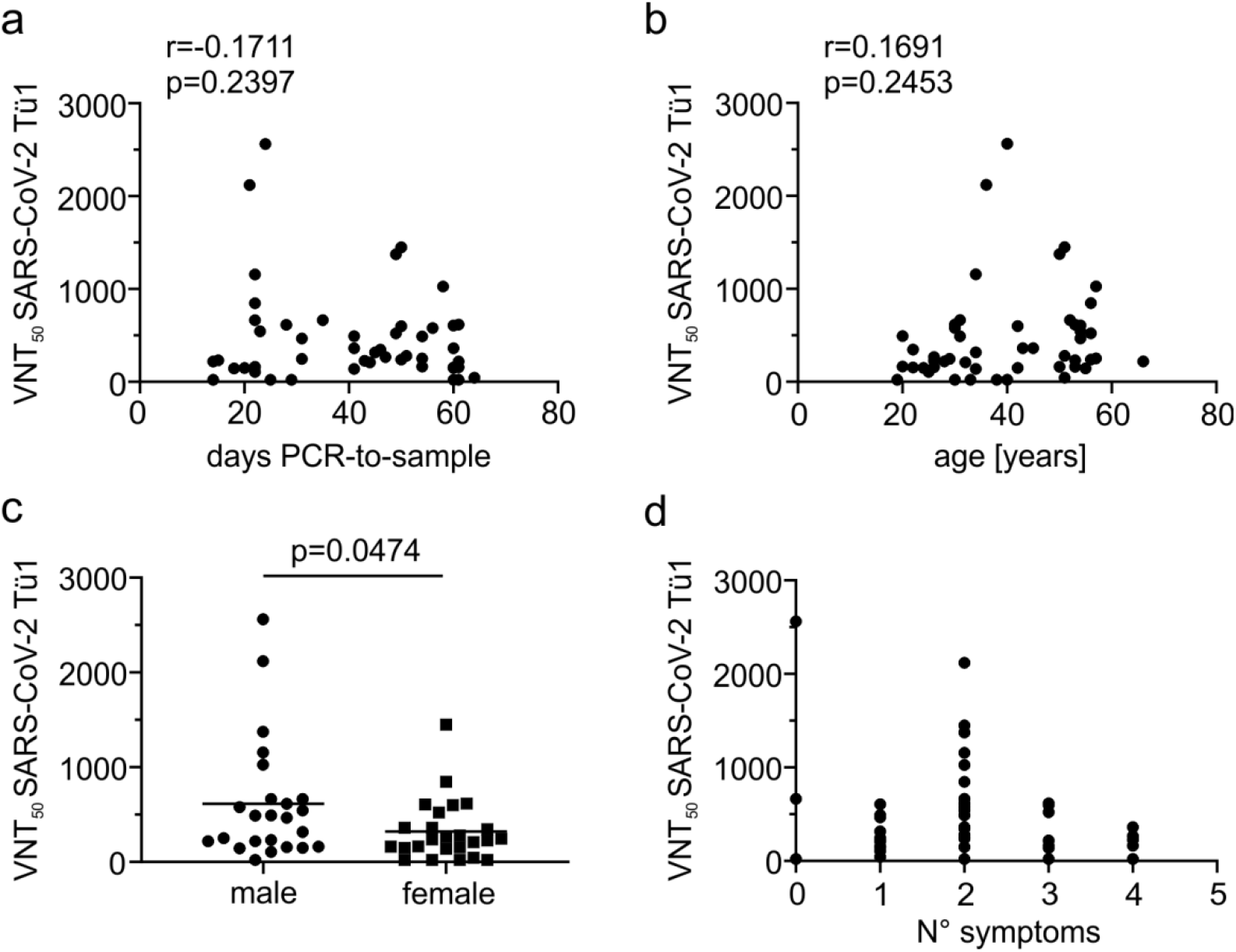
Association of patient characteristics with sera VNT_50_. The VNT_50_ of each patient serum was associated with the individual (a) date of the positive SARS-CoV-2 qRT-PCR diagnostic test to blood sampling, (b) the age of the patient, (c) the gender and (d) the number of symptoms reported. Statistical analyses were done with an unpaired two-tailed student’s t-test. See detailed patient characteristic in Supplementary Table 1.

### SARS-CoV-2-specific IgG against the S-protein RBD indicate serum neutralization

Next, we set out to define serological correlates of virus neutralization *in vitro*. Overall, the neutralizing capacity of the sera correlated with the abundance of SARS-CoV-2 specific IgG against the S-protein (r=0.6137; Fig. 3a), with a slightly better r-value when the IgG-measured were RBD-specific (r=0.7198; Fig. 3b). This indicates, as expected, that antibodies against the RBD are nvolved in SARS-CoV-2-neutralization. Similarly, RBD-specific IgA and IgM correlated with neutralization (Fig. 3c and Fig. 3d), even though their abundance is highly diverse in the patient cohort (Supplementary Table 1). In contrast, IgGs against the SARS-CoV-2-nucleocapsid measured by the Roche ECLIA poorly correlated with serum neutralization (r=0.3249; Fig. 3d).

**Figure 3.**
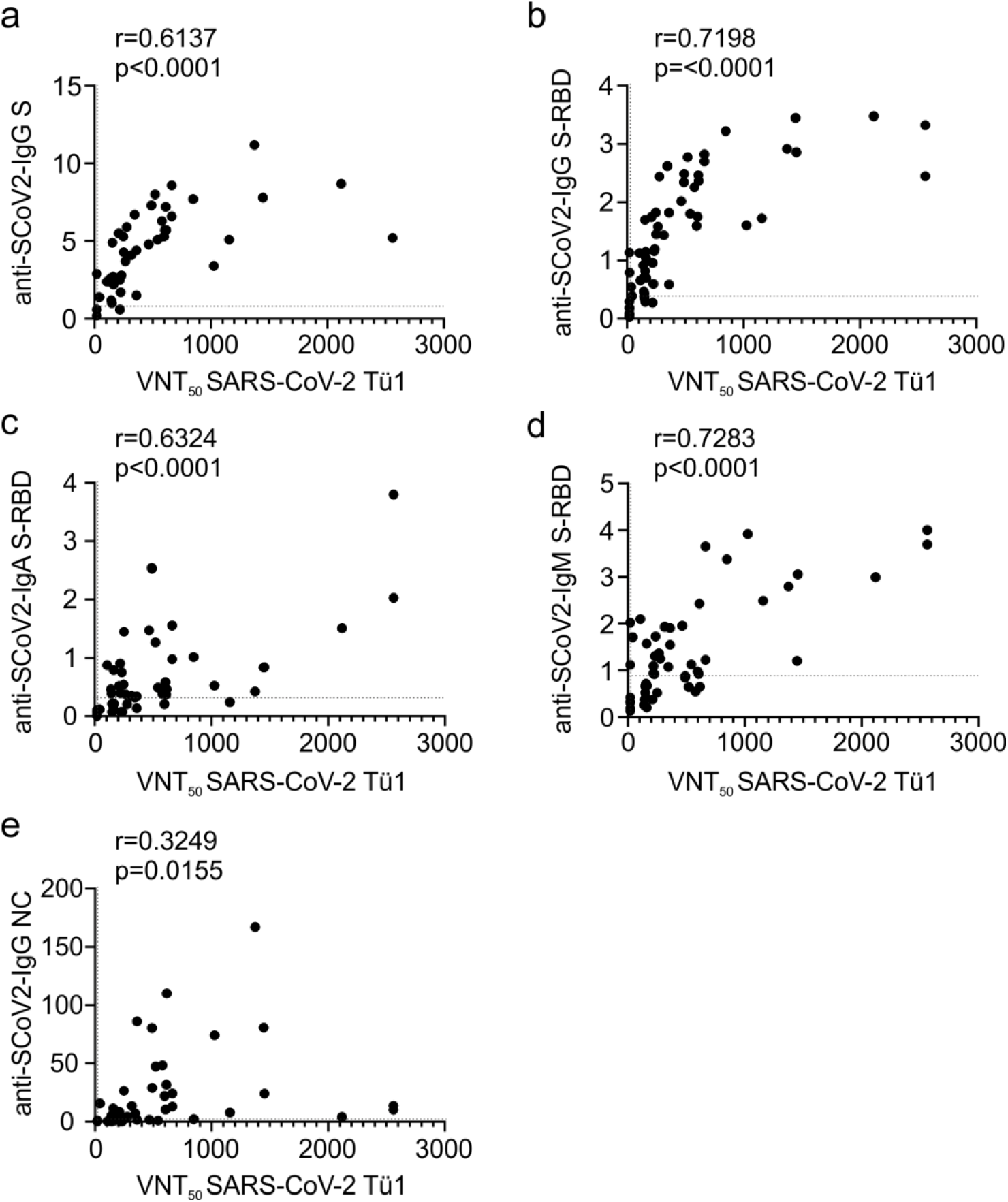
Correlation of serological parameters with sera VNT_50_. The VNT_50_ of each patient serum was correlated with (a) the value of SARS-CoV-2-S-specific IgGs measured by the Euroimmune ELISA, (b) the relative quantitative value of SARS-CoV-2-S-RBD-specific IgGs measured by the Mediagnost ELISA, (c) the relative quantitative value of SARS-CoV-2-S-RBD-specific IgAs measured by the Mediagnost ELISA, (d) the relative quantitative value of SARS-CoV-2-S-RBD-specific IgMs measured by the Mediagnost ELISA (e) the relative quantitative value of SARS-CoV-2-NC-specific IgGs measured by the Roche ECLIA. Dotted lines indicate the respective assay thresholds

In conclusion, ELISAs or antibody tests, quantifying antibodies against the S-protein and in particular the S-RBD correlate best with patient serum neutralization.

### Antibodies against seasonal coronavirus 229E correlate with serum neutralization of SARS-CoV-2

It is a matter of ongoing debate if antibodies against seasonal coronaviruses might confer cross-protection against SARS-CoV-2. To get first insights into this question, we took use of a quantitative throughput Western blot-based detection system identifying the bulk of IgGs against a specific coronavirus (16). As expected and in line with our previous data (Fig. 3a and 3b), IgG against SARS-CoV-2 correlated with VNT_50_ (r=0.6592; Fig. 4a). Of note, IgG against the seasonal coronavirus 229E was also associated with VNT_50_ (r=0.4136, p=0.0017; Fig. 4b), indicating that this class of antibodies might support SARS-CoV-2 neutralization. Remarkably, this effect was specific for 229E and neither observed for seasonal coronaviruses OC43 (Fig. 4c) nor NL63 (Fig. 4d). In conclusion, even though based on correlation analyses, our data indicates that a humoral immune response against the seasonal coronavirus 229E might support SARS-CoV-2 neutralization.

**Figure 4.**
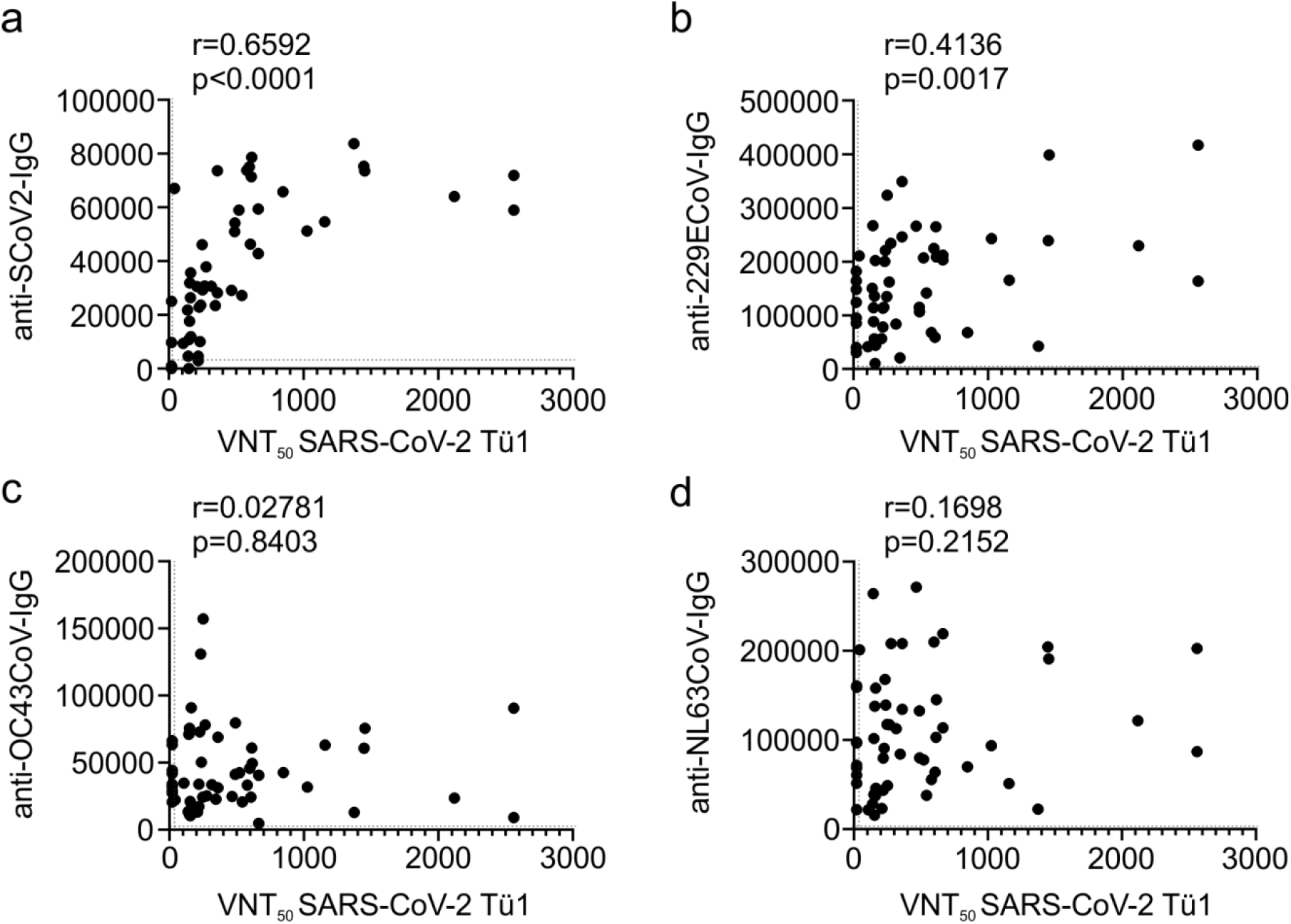
Correlation of antibodies against seasonal coronaviruses with sera VNT_50_. The VNT_50_ of each patient serum was correlated with the relative quantitative value of a throughput diagnostic Western blot detection system measuring CoV-specific IgG against (a) SARS-CoV-2, (b) CoV-229E, (c) CoV-OC43 or (d) CoV-NL64. Dotted lines indicate the respective assay thresholds defined as positive.

## Discussion

Recent studies assessed the development of virus-specific antibodies in various cohorts of COVID-19 convalescent individuals (12, 13, 19-21). Overall, the data of the latter is in accordance with ours, showing that the vast majority of individuals develop SARS-CoV-2-specific antibodies. Furthermore, SARS-CoV-2-specific IgGs were more prevalent than IgMs (20, 21), a finding that we confirm and extend to the abundance of SARS-CoV-2-specific IgA. Using pseudovirus-based neutralization assays Robbiani et al. (20) also report that there is a correlation between VNT_50_ and antibodies against the S-RBD. Furthermore, males had significantly higher neutralizing activity than females, a finding which is also supported by our data (Fig. 2c). We extend the aforementioned studies by several aspects. First, for all our neutralization experiments we employed a fully infectious clinical SARS-CoV-2-isolate on a human cell line. Second, we performed a comprehensive comparison of several serological tests to delineate correlates of SARS-CoV-2 neutralization. This revealed that NC-specific antibodies poorly correlate with serum virus neutralization. In contrast, as supported by the findings of Robbiani et al. (22) and Ju et al (23) RBD-specific IgGs correlate best with serum neutralization (Fig. 3b). In this context, it is noteworthy that S-RBD specific IgA and IgM also showed a high degree of correlation with the VNT_50_ (Fig. 3c and 3d), indicating that these antibodies, even though their abundance was highly diverse in our patient cohort, might contribute to serum neutralization.

A phenomenon that is critically discussed is the potential enhancement of infection by non-neutralizing antibodies (ADE) (14). For our VNT-assays, we are using human cells expressing a diverse set of F_c_-receptors, and directly assess the rate of infected cells by immunofluorescence or reporter-gene expression. Hence, we should be able to detect enhancement of infection by serum that is not or only poorly neutralizing and in higher dilution ranges. However, in none of our 49 sera we observed ADE at any of the dilutions tested, indicating that at least antibodies generated in the natural context of SARS-CoV-2 infection do not contribute to severity of infection. This is in line with the absence of a correlation between the number of symptoms and VNT_50_ in our patients (Fig. 2d). On the other hand, our cohort is biased due to the fact that none of the patients was hospitalized. Therefore, it will be important to analyze if ADE plays a potential role in severe cases of COVID-19.

Up to now, it was elusive if antibodies against seasonal coronaviruses that are highly prevalent within the human population play a role in SARS-CoV-2 neutralization. We employed an innovative throughput Western blot system to concomitantly detect antibodies specific against SARS-CoV-2 as well as the seasonal coronaviruses 229E, OC43 and NL63 (16). In fact, 100% of individuals included in our study had antibodies against the three seasonal coronaviruses with high diversity in relative numbers (Supplementary Table 1). Correlating the latter with our VNT_50_ values revealed a significant association of 229E-specific IgGs with the ability of patient sera to neutralize SARS-CoV-2 infection (Fig. 4b). This effect was 229E-specific since none of the other seasonal coronaviruses showed such an association (Fig. 4c and 4d). While it is clear that 229E-specific IgGs are not sufficient to confer cross-protection against SARS-CoV-2, our data imply, based on correlation analyses, that the prevalence of such antibodies might assist in the neutralization of SARS-CoV-2. A hypothesis that is in line with the observation that antibodies directed against the RBD of SARS or MERS alone are not sufficient to inactive SARS-CoV-2 (23). It will be highly interesting to analyze if the epidemiology of seasonal coronaviruses is a determinant of COVID-19 severity, with the implication that in areas with a high prevalence of antibodies against 229E mortality is decreased.

Altogether, we here establish several correlates of SARS-CoV-2 neutralization by patient serum using a relevant virus-neutralization test. Even though S-RBD-specific IgGs correlate best with serum neutralization, it is clear that multiple factors contribute to a potent neutralizing antibody response. This might include subclasses of S-specific antibodies as for instance IgM and IgA as well as the antibody response elicited against the seasonal coronavirus 229E. This makes it particularly difficult to define singular serological correlates of immune protection as discussed in the context of COVID-19 “immunity passports”. Furthermore, such an approach neglects other potentially essential factors of immune protection as for instance, T-cell mediated immunity (24, 25) and the innate immune response (26).

## Data Availability

All relevant data is within the manuscript and the supplementary information file.

## Author contributions

NR, RB, and MS established the neutralization test and performed all experiments with SARS-CoV-2; KA and TB provided patient samples and did the Euroimmune ELISA; BF performed all ELISAs provided by Mediagnost; KH established, analyzed and supervised the Roche ECLIA; SF, FR and MT established, conducted and analyzed the DigiWest detection system for SARS-CoV-2 and seasonal CoVs; NR, RB, and MS analyzed the data; NR and MS drafted the figures and wrote the manuscript. MS developed the manuscript to its final form; MS planned and supervised the study; all authors read, edited, and approved the final manuscript.

## Acknowledgements

We thank all patients and healthy volunteer donors for their cooperation in this study and for their swab specimen collection and blood donation. We thank Thomas Iftner and Tina Ganzenmüller for their support with diagnostic tests and the collection of swab specimens.

## Methods

### Study participants and sample processing

Blood was drawn from potential blood donors for reconvalescent plasma therapy after written consent at the Clinical Transfusion Medicine, Tübingen between April 04 and May 12, 2020, under the guidelines of the local ethics committees 222/2020BO. All patients (n=49) were older than 18 years old and provided a PCR-confirmed diagnosis of SARS-CoV-2 (n=46) and three were symptomatic and close contacts to positively diagnosed COVID-19 patients (partners tested positive). All patients were non-hospitalized with asymptomatic to mild courses of disease and they were fully convalescent showing no symptoms on the day of blood donation. Basic demographic information was collected including age and sex, as well as self-perceived symptoms (cough, fever, limb pain and headache, diarrhea, and loss of taste). In addition, blood from four healthy donors and one hospitalized patient was collected (Supplementary Table 1). Serum samples were stored at −80°C.

### Cell culture

Caco-2 (Human Colorectal adenocarcinoma) cells were cultured at 37 °C with 5% CO2 in DMEM containing 10% FCS, with 2 mM l-glutamine, 100 μg/ml penicillin-streptomycin and 1% NEAA.

### Viruses

A throat swab sample collected in March 2020 at the diagnostic department of the Institute for Medical Virology and Epidemiology of Viral Diseases, University Hospital Tübingen, from a SARS-CoV-2 positive patient was used to isolate the virus (200325_Tü1). 50 µl of patient material was diluted in media, sterile-filtrated, and used directly to inoculate 200.000 Caco-2 cells in a 6-well. 48 hpi (hours post-infection) the supernatant was collected, centrifuged, and stored at −80°C. Supernatant as well as cell lysates from infected cells were tested by Western blot using a SARS-CoV-2 anti-nucleocapsid protein (NP) specific antibody (GeneTex). In addition, the identity of the virus was confirmed by qRT-PCR diagnostics via S and E gene amplification. An aliquot of the isolate was used to amplify the virus in a medium flask of Caco-2 cells (2X10^6^ cells) in 13 ml DMEM + supplements and 5% FCS. 48 hpi, the supernatant was centrifuged and stored in aliquots at −80°C.

The recombinant SARS-CoV-2 expressing mNeonGreen (icSARS-CoV-2-mNG) (22) was obtained from the World Reference Center for Emerging Viruses and Arboviruses (WRCEVA) at the UTMB (University of Texas Medical Branch). To generate icSARS-CoV-2-mNG stocks, Caco-2 cells were infected as above, the supernatant was harvested 48 hpi, centrifuged, and stored at −80°C.

For MOI determination, a titration using serial dilutions of both virus stocks (200325_Tü1 and mNG) was conducted. The number of infectious virus particles per ml was calculated as the (MOI x cell number)/(infection volume), where MOI = -ln(1 - infection rate). To reach an infection rate of ~20% an MOI of 0.3 was used for SARS-CoV-2-200325_Tü1 and 1.1 for SARS-CoV-2.mNG.

**Enzyme-linked Immunosorbent Assays (ELISAs); Euroimmun**, the Euroimmun SARS-CoV-2-ELISA (IgG) (Euroimmun, Lübeck, Germany) with the recombinant S1 target antigen of SARS-CoV-2 was performed according to manufacturer’s instructions in serum. Patient samples are diluted 1:101 in sample buffer. The included controls and calibrator in the test kit were used with each run. Results are given as ratios (optical density (OD) of control or clinical sample/OD of calibrator). According to the manufacture, ratios were classified as negative (< 0.8), borderline (≥ 0.8 - < 1.1) and positive (≥ 1.1). **Mediagnost**, IgG antibody detection directed to the S1 RBD SARS-CoV-2 in human sera using Mediagnost test system was made according to the manufacturer’s instructions. Briefly, these tests are two-step enzyme-linked immunosorbent assays. The solid phase consists of a 96-well Microtiter plate (Greiner, Bio-One, Frickenhausen Germany) that is coated with the recombinant SARS-CoV-2 spike protein S1. The antibodies from patients that are directed against SARS-CoV-2 S1 protein bind to the solid phase coated S1 protein. Next a horseradish peroxidase (HRP) conjugated goat anti-human IgG binds to the human IgG antibodies. The following step involves the substrate for the HRP being added by which the substrate is converted from colorless into blue color; and after addition of a stop solution the color changes to yellow. The extinction of the yellow solution can be measured at a wavelength at 450 nm with reference at 620 nm. Increasing extinctions represent increasing amounts of antibodies to SARS-CoV-2 S1 protein. Samples showing extinctions that were three times higher than the negative control can be interpreted as being positive for anti-SARS-CoV-2 S1. IgA and IgM antibody detection directed to the SARS-CoV-2 S1 protein was made in analogy to the above-described IgG detection system except that the HRP labeled detection antibody was directed against human IgA or human IgM antibodies. According to the manufacture, ratios were classified as negative (<0,42), borderline (≥0,42-0,7) and positive (≥0,7) for IgG, (<0,33), (≥0,33-0,7) and (≥0,7) for IgA and (<0,87), (≥0,87-1,47) and (≥1,47) for IgM.

### Elecsys anti-SARS-CoV-2 (Roche)

For qualitative detection of anti SARS-CoV-2 (IgG+IgM) antibodies the electrochemiluminescence immunoassay (ECLIA) was performed using the fully automated cobas e 6000/601 immunoassay analyzer (Roche Diagnostics, Mannheim, Germany). This assay targets recombinant SARS-CoV-2 nucleocapsid (NC) protein. Two calibrators are used (Cal1 nonreactive, COI 0,101; and Cal2 reactive, COI 1,2) in the double antigen-sandwich based assay (SARS-CoV -2 recNC biotin label, and SARS-CoV-2 recNC ruthenium complex label). Each, 20μl of sera and reference solutions were used, immune complexes are fixed to streptavidin-coated microparticles. Read out is given in relative light units in the form of cutoff index (COI, signal/cutoff). The Elecsys reagents derived from LOT 49500101. For negative control (<150% Cal1), we used pooled sera from 100 mothers at birth of the Tuebingen congenital CMV study 2012. Furthermore, we used a negative control serum from a direct Covid-19 contact person, repeatedly negative tested for SARS-CoV-2 RNA and nucleocapsid-specific antibodies without any symptoms during and after a one week close exposition. For positive control, we used a dilution series of a serum from a reconvalescent student infected symptomatically (fever, cough, loss of smell) tested posiive for viral RNA and NC-specific antibodies. The COIs ranged from 100 to 1. If the numeric COI result was ≥ 1,0, the serum was diagnosed as reactive, COI <1,0 were attributed as non-reactive. COI values of the positive controls were stable over at least 2 months.

### Multiplexed detection of anti-coronavirus antibodies

Whole viral protein lysates from 229E, OC43, and NL63 (ZeptoMetrix Corp) and from SARS-CoV-2 were used for DigiWest as described (16). Viral protein lysates were used for denaturing gel electrophoresis and Western blotting using the NuPAGE system. Blot membranes were washed with PBST (0.1% Tween-20, PBS) and membrane-bound proteins were biotinylated by adding 50 μM NHS-PEG12-Biotin (Thermo Fisher Scientific) in PBST for 1 h. After washing in PBST, membranes were dried overnight. Subsequently, the Western-Blot lanes were cut into 96 strips of 0.5 mm width and were transferred to a 96-well plate (Greiner Bio-One). For protein elution, 10 µL of elution buffer was added to each well (8 M urea, 1% Triton-X100 in 100 mM Tris-HCl pH 9.5). The protein eluates were diluted with 90 µL dilution buffer (5% BSA in PBST, 0.02% sodium azide).

Neutravidin-coated MagPlex beads (Luminex) of a distinct colour ID were added to the protein eluates and binding was allowed overnight; 500 μM PEG12-biotin in PBST was added to block remaining Neutravidin binding sites. The bead containing fractions were pooled and thereby the original Western blot lanes were reconstituted. Beads were washed in PBST and resuspended in store buffer (1% BSA, 0.05 % azide, PBS). The generated bead-set represents the proteomes of the four coronaviruses (SARS-CoV-2, OC43, 229E, NL63) and reactivity against all proteins can be tested in one assay.

For serum incubation, 5 µL of the bead mix were equilibrated in 50 µL serum assay buffer (Blocking Reagent for ELISA (Roche) supplemented with 0.2% milk powder, 0.05% Tween-20 and 0.02% sodium azide, 25% Low Cross buffer (Candor Bioscience), 25% IgM-reducing agent buffer (ImmunoChemistry). Serum assay buffer was discarded and 30 µL of diluted patient serum (1:200 in serum assay buffer) was added and incubated for 2 hours at RT on a shaker. After washing in PBST, 30 µL of Phycoerythrin labelled anti-human IgG secondary antibody (diluted 1:200 in serum assay buffer; Dianova) was added and incubated for 45 min at 23 °C. The beads were washed twice with PBST and readout was performed on a Luminex FlexMAP 3D.

The DigiWest analysis tool was used to assess serum reactivity against the viral proteins (16). Virus protein-specific peaks were identified and average fluorescence intensity (AFI) values were calculated by integration of peak areas.

### Neutralization assay

For neutralization experiments, 1×10^4^ Caco-2 cells/well were seeded in 96-well plates the day before infection in media containing 5% FCS. Cells were co-incubated with SARS-CoV-2 clinical isolate 200325_Tü1 at a MOI=0.3 and patient sera in serial two-fold dilutions from 1:20 up to 1:2560. 48 hpi cells were fixed with 80% acetone for 5 minutes, washed with PBS, and blocked for 30 minutes at room temperature (rt) with 10 % normal goat serum (NGS). Cells were incubated for 1 h at rt with 100 µl of serum from a hospitalized convalescent donor in a 1:1000 dilution and washed 3 times with PBS. 100 µl of goat anti-human Alexa594 1:2000 in PBS was used as secondary antibody for 1h at rt. Cells were washed 3 times with PBS and counter-stained with 1:20000 DAPI solution (2 mg/ml) for 10 minutes at rt. For quantification of infection rates images were taken with the Cytation3 (Biotek) and DAPI+ and Alexa594+ cells were automatically counted by the Gen5 Software (Biotek).

Alternatively, Caco-2 cells were co-incubated with the SARS-CoV-2 strain icSARS-CoV-2-mNG at a MOI=1.1 and patient sera in serial two-fold dilutions from 1:40 up to 1:5120. 48 hpi cells were fixed with 2% PFA and stained with Hoechst33342 (1 μg/mL final concentration) for 10 minutes at 37°C. The staining solution was removed and exchanged for PBS. For quantification of infection rates images were taken with the Cytation3 (Biotek) and Hoechst+ and mNG+ cells were automatically counted by the Gen5 Software (Biotek). Virus neutralizing titers (VNT_50_) were calculated as the half-maximal inhibitory dose (ID50) using 4-parameter nonlinear regression (GraphPad Prism).

### Software and statistical analysis

GraphPad Prism 8.0 was used for statistical and correlation analyses and to generate graphs. Figures were generated with CorelDrawX7. Other software used included Gen5 v.3.04.

